# An algorithmic procedure for measuring deep brain stimulation-induced capsular activation using motor evoked potentials

**DOI:** 10.1101/2025.08.26.25334489

**Authors:** Eric R. Cole, Enrico Opri, Seyyed Bahram Borgheai, Yuji Han, Faical Isbaine, Nicholas Boulis, Jon T. Willie, Nicholas AuYong, Robert E. Gross, Svjetlana Miocinovic

**Author notes:** **Corresponding Author** Svjetlana Miocinovic, MD, PhD, Department of Neurology, Emory School of Medicine, Atlanta, GA, USA 30322.

## Abstract

**Objective:** Effective deep brain stimulation (DBS) treatment for Parkinson’s disease requires careful adjustment of stimulation parameters and targeting to avoid motor side effects caused by activation of the internal capsule. Currently, patients must self-report side effects during device programming and implantation surgery – a challenging and subjective process that could lead to suboptimal therapy or exacerbate the time needed to optimize treatment. Motor evoked potentials (mEP), the use of electromyography to record DBS-induced muscle activation, offer a promising biomarker for objective motor side effect detection.

**Approach:** Here, we present an automated algorithmic procedure for mEP detection and quantification.

**Main Results:** First, we design and evaluate a series of signal processing techniques to accurately detect mEP while mitigating the influence of stimulation artifacts and noise, then demonstrate a strategy for integrating multi-channel EMG responses into a single side effect biomarker (the mEP score). Next, we use data from a large patient cohort of intraoperative recordings (N = 54 STN leads) to quantify several physiological features of mEP, including their response frequency, latency, amplitude, and waveform similarity properties. Last, we show that the mEP score responds to DBS amplitude and contact configuration parameters in a manner that is consistent with expected STN-capsular anatomy.

**Significance:** The results of this study inform an end-to-end approach for side effect biomarker measurement that could aid the precision and efficiency of DBS programming and surgical targeting.

## 1. Introduction

Deep brain stimulation (DBS) is a transformative treatment for movement disorders, including Parkinson’s disease, dystonia, essential tremor, and more.^1^ However, DBS can often produce undesirable side effects which arise from activation of pathways adjacent to the brain region targeted for therapeutic benefit. Assessing side effects is crucial during two clinical procedures: first, during surgery, intraoperative side effect assessment helps guide and confirm accurate electrode placement within the brain, avoiding undesirable sites.^2^ Second, clinicians must program DBS parameters postoperatively to maximize therapeutic benefit while minimizing side effects^3^ – a burdensome trial-and-error process that typically requires more than a dozen hours of labor per patient.^4^

One critical challenge in providing effective DBS therapy comes from subjectivity and lack of standardization in measuring side effects.^5^ Such side effects can include unwanted contraction of facial and limb muscles (here referred as motor side effects),^6^ as well as paresthesia,^7^ dyskinesia,^8^ mood changes,^9^ and more.^10^ Motor and sensory side effects occur when DBS is delivered at an excessively high amplitude, causing activation of white matter pathways that are adjacent to the desired stimulation target, including the corticospinal and corticobulbar tracts of the internal capsule (motor) and the medial lemniscus (sensory).^3^ Currently, there is no standardized scoring system for quantifying DBS-induced side effects, as they can express in many semiological forms and express with high variability (e.g. at different locations or sensations) across patients. Instead, the current standard of care relies on patients to self-report their side effects or clinicians to visually observe stimulation induced muscle contractions, a process that is challenging and often unreliable. Specifically, patients can over-report perceived side effects, due to difficulty in differentiating DBS-induced muscle contractions from disease symptoms such as rigidity, dystonia, or pain.^11^ Similarly, patients may under-report side effects if capsular activation is subtle or cognitive dysfunction or anxiety limit their active participation during surgery or programming. Finally, side effects measurement is challenging when patients are anesthetized during surgery. The unreliability of subjective side effect evaluation can lead to suboptimal programming outcomes or exacerbate the length of time needed to find useful settings.^12^

A second major challenge in providing effective DBS comes from the increasing resolution of current and next-generation DBS hardware. A DBS setting is defined by the amplitude, frequency, and pulse width of electrical current, as well as the contact configuration at which current is delivered. First-generation DBS devices featuring 4 contacts could enable 16 different permutations of stimulating contacts. Modern 8-contact devices and recently introduced 16-contact devices raise the number of possible parameter permutations into the millions.^13,14^ Higher-resolution DBS devices could improve patient outcomes by providing more precise ability to target specific pathways and symptoms; however, they will very quickly prove nearly impossible to program manually.^15^ A biomarker of induced side effects could therefore improve the use of DBS by making side effect measurement objective and standardized, providing quickly measurable feedback to make programming more efficient, and better enabling the practical use of high-resolution devices.

While multiple biomarkers have shown promise in predicting symptom relief with DBS, biomarkers of DBS-induced side effects are equally important but have received comparatively little attention in the literature.^16^ One promising biomarker is the motor evoked potential (mEP), the use of electromyography (EMG) to measure DBS-induced muscle activation, which can objectively quantify motor side effects caused by DBS-induced capsular activation. mEP have been shown to predict the amplitude thresholds at which different DBS settings induce patient-reported motor side effects^17^ and correlate with electrode distance to the internal capsule.^18^ mEP could therefore provide a biomarker that can be quickly measured (within seconds) to provide objective feedback for DBS programming and surgical targeting. However, prior mEP studies have primarily used blinded manual inspection of time-series data to identify which stimulation settings induced mEP;^18,19^ there remains a need for robust algorithmic tools to automatically measure mEP and distinguish physiological response from various sources of noise that can arise in EMG recordings. Such tools can aid further investigation into the clinical utility of mEP, particularly with large-scale data collected from many patients, and will be necessary for its use in closed-loop and real-time clinical applications. Notably, in this study we focus only on mEP as a biomarker for DBS motor side effects; while DBS can cause other side effects, these often habituate or can be addressed by medication changes,^20^ whereas motor side effects present the most therapy-limiting side effect for effective DBS.

In this study, we develop an algorithmic procedure for using motor evoked potentials to provide a fast, robust, and objective biomarker that could be used to quantify whether a given DBS setting induces activation of the internal capsule and validate its relationship to capsular anatomy within our patient cohort. First, we evaluate a wide array of signal processing and stimulation artifact rejection techniques to develop an accurate, automated algorithm for detecting mEP. Second, we demonstrate a strategy for normalizing and integrating detected mEP response amplitudes across different recorded muscles into a single interpretable value (the “mEP score”) to quantify capsular activation for a given setting. Next, we use recordings collected from a large cohort of patients undergoing DBS implantation surgery in the subthalamic nucleus (STN) (N = 54 leads) to show that several muscle-specific physiological features of mEP, including latency and response variability, reflect capsular anatomy. Last, we show that the mEP score responds to DBS amplitude and contact configuration parameters in a manner that is consistent with conventional observations of DBS-induced side effects and expected STN-capsular anatomy.

## 2. Methods

### 2.1 Experiment protocol for EMG recording during DBS implantation surgery

Patients with idiopathic Parkinson’s disease scheduled to undergo awake STN DBS surgery were recruited for the study. Written informed consent was obtained before surgery under the protocol approved by the Institutional Review Board. DBS electrodes (both quadripolar and directional leads: Medtronic 3389, Medtronic B33005, Boston Scientific Vercise Cartesia Directional Lead 2202, and Abbott Infinity 6172) were implanted using microelectrode guidance and standard clinical procedures. A 2”x4” pad electrode was placed on the shoulder ipsilateral to the stimulated hemisphere to provide the return path for monopolar stimulation. EMG electrodes (Natus Medical EMG disposable dual electrodes were used for limb muscles; Natus Medical EMG disposable disk electrodes were used for facial muscles due to smaller size and lighter weight) were placed on up to 8 muscles per patient (Figure 1b; listed in Table 1), including the contralateral arm: biceps, flexor carpi radialis (FCR), extensor carpi radialis (ECR), and first dorsal interosseous (FDI); and bilateral face: nasalis, orbicularis oris (orb oris), genioglossus. We did not record from lower extremity muscles after observing no response in a small initial patient cohort. The DBS electrode, connected to the NeuroOmega clinical electrophysiology system (Alpha Omega Engineering) with custom connectors, was used for recordings and to deliver stimulation. Recordings were performed at least 12 hours after stopping all anti-parkinsonian medications and at least 30 minutes after stopping propofol sedation. EMG signals were recorded at 22 kHz sampling rate with built-in hardware bandpass filtering between 0.075 and 3500 Hz. EMG signals were recorded from bipolar-referenced electrode pairs with a common ground on the contralateral knee (Kendall 535 hydrogel ECG electrode).

**Figure 1.**
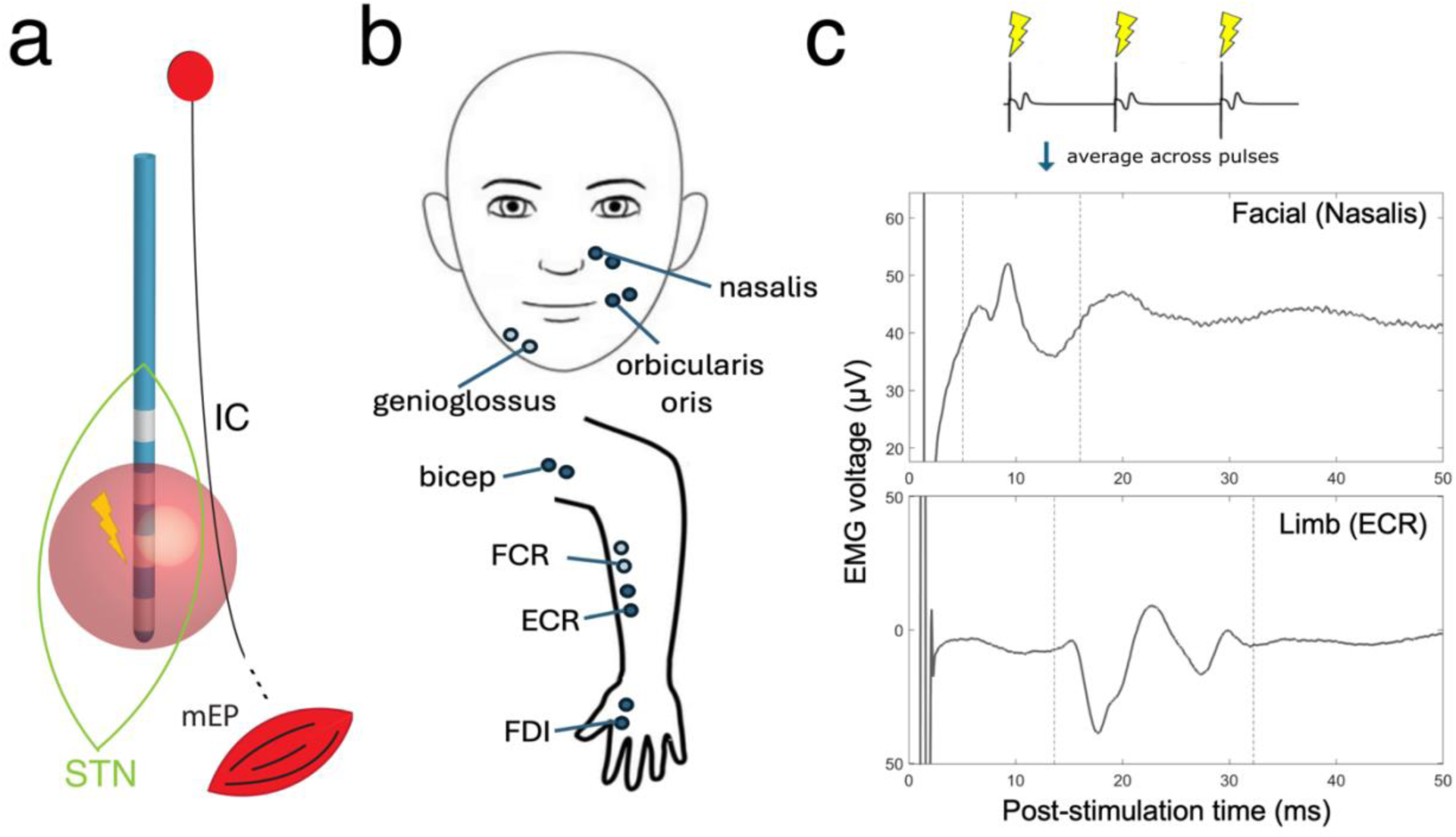
Overview of experiment design for intraoperative deep brain stimulation and electromyography. a) Circuit diagram of deep brain stimulation in the subthalamic nucleus and adjacent structures. Blue: implanted deep brain stimulation lead. Green: anatomical outline of the subthalamic nucleus. IC: internal capsule. Red sphere: hypothetical volume of tissue activated by the DBS-generated electric field, partially overlapping with the internal capsule. Bottom red shell: activated muscle tissue. b) Diagram of EMG bipolar electrode placement for commonly recorded muscles in this study. ECR: extensor carpi radialis. FCR: flexor carpi radialis (transparent marker indicating placement underneath the anatomical structure). FDI: first dorsal interosseus. c) Example pulse-averaged EMG recordings displaying a motor evoked potential response in nasalis (top) at 10 ms latency and ECR (bottom) at 18 ms latency.

**Table 1.** EMG experiment data by muscle. N. Leads: the number of DBS leads for which that muscle was recorded. N. Settings: the total number of stimulation settings during which that muscle was recorded. Pct. Settings Excluded: the percentage of times, when recording from the given muscle, a stimulation setting was excluded based on signal exclusion criteria. Detection window: the time window used to guide peak detection for the given muscle. IL: ipsilateral. CL: contralateral. For very infrequently recorded facial muscles (genioglossus), the data-derived latency window obtained for nasalis was used instead.

|  | Muscle | N. Leads | N. Settings | Pct. Settings Excluded (%) | Detection window (ms) |
| --- | --- | --- | --- | --- | --- |
| Limb Muscles | CL ECR | 53 | 1434 | 12.3 | 16.5-30.5 |
|  | CL FCR | 41 | 1178 | 13.2 | 14.5-29.5 |
|  | CL FDI | 41 | 1249 | 12.6 | 21.0-30.0 |
|  | CL Bicep | 17 | 763 | 11.4 | 11-29.0 |
| Facial Muscles | CL Genioglossus | 7 | 277 | 22.4 | 5.0-18.0 |
|  | CL Nasalis | 38 | 1169 | 10.9 | 5.0-18.0 |
|  | CL Orb. Oris | 31 | 935 | 14.5 | 5.0-18.5 |
|  | IL Genioglossus | 2 | 80 | 0 | 5.0-18.0 |
|  | IL Nasalis | 21 | 734 | 16.9 | 5.0-18.0 |
|  | IL Orb. Oris | 15 | 500 | 22.2 | 5.0-18.5 |

### 2.2 Stimulation delivery and parameters

Stimulation was applied for 12 seconds with 3 seconds pause between settings, using biphasic pulses delivered at 10-Hz frequency to allow sufficient time to measure the mEP waveform based on its post-stimulation peak latency. Stimulation settings were delivered in randomized order at a wide variety of different amplitude values (typically 1-5 mA), pulse width values (30-120 microseconds), and both monopolar (ring, pseudoring or segments) and bipolar contact configurations. The stimulation waveform was asymmetric biphasic with 70 µs inter-phase duration and the first phase at higher amplitude, designed to approximate a conventional clinical DBS waveform. For most stimulation settings (60 µs or less), the second phase was 8 times longer with 1/8 of the amplitude of the first phase to maintain charge balance between phases. Because the Neuro Omega system implements a maximum 500 µs limit for each phase duration, stimulation settings with pulse widths longer than 60 us had their second phase duration limited to 480 µs and second phase amplitude adjusted accordingly to maintain charge balance. Total number of stimulation settings varied per patient (mean 24 ± 17 settings) as allowed given intraoperative time limitations.

To normalize encoding of the DBS parameter space across manufacturers, which use different channel ordering schemes and different nomenclature to refer to the device contact orientation, we standardized the stimulation parameter encoding for each lead to follow a geometric naming scheme throughout the results. The 4 device levels are numbered 1 (ventral contact) – 4 (dorsal contact). The segments are named based on their anatomical orientation: A (positioned anteriorly during standard surgical practice), M (postero-medial segment), and L (postero-lateral segment).

### 2.3 Motor evoked potential detection

Various sources of noise, electrode placement, and artifacts common to surface EMG recordings make it non-trivial to algorithmically detect mEPs. First, the short peak latency of facial muscles can overlap in time with post-stimulation amplifier recovery artifacts (shown in Fig. 2b); thus, stimulation artifacts can produce false positive detections, and stimulation artifact removal could particularly improve facial muscle detection. Additionally, various sources of noise (low-frequency fluctuations e.g. due to cardiac activity, tremor, voluntary movement or breathing motion; high-frequency noise from nearby electronics; 60-Hz line noise and its harmonics) can produce both false positive (introducing detectable peaks) and false negative detections (by corrupting baseline calculations and leading to prominent responses remaining undetected). Below, we first describe a peak detection strategy for quantifying whether a given stimulation setting produced an mEP response. Next, we describe several different artifact rejection and filtering steps we evaluated alongside peak detection to combat the above difficulties and improve detection performance.

**Figure 2.**
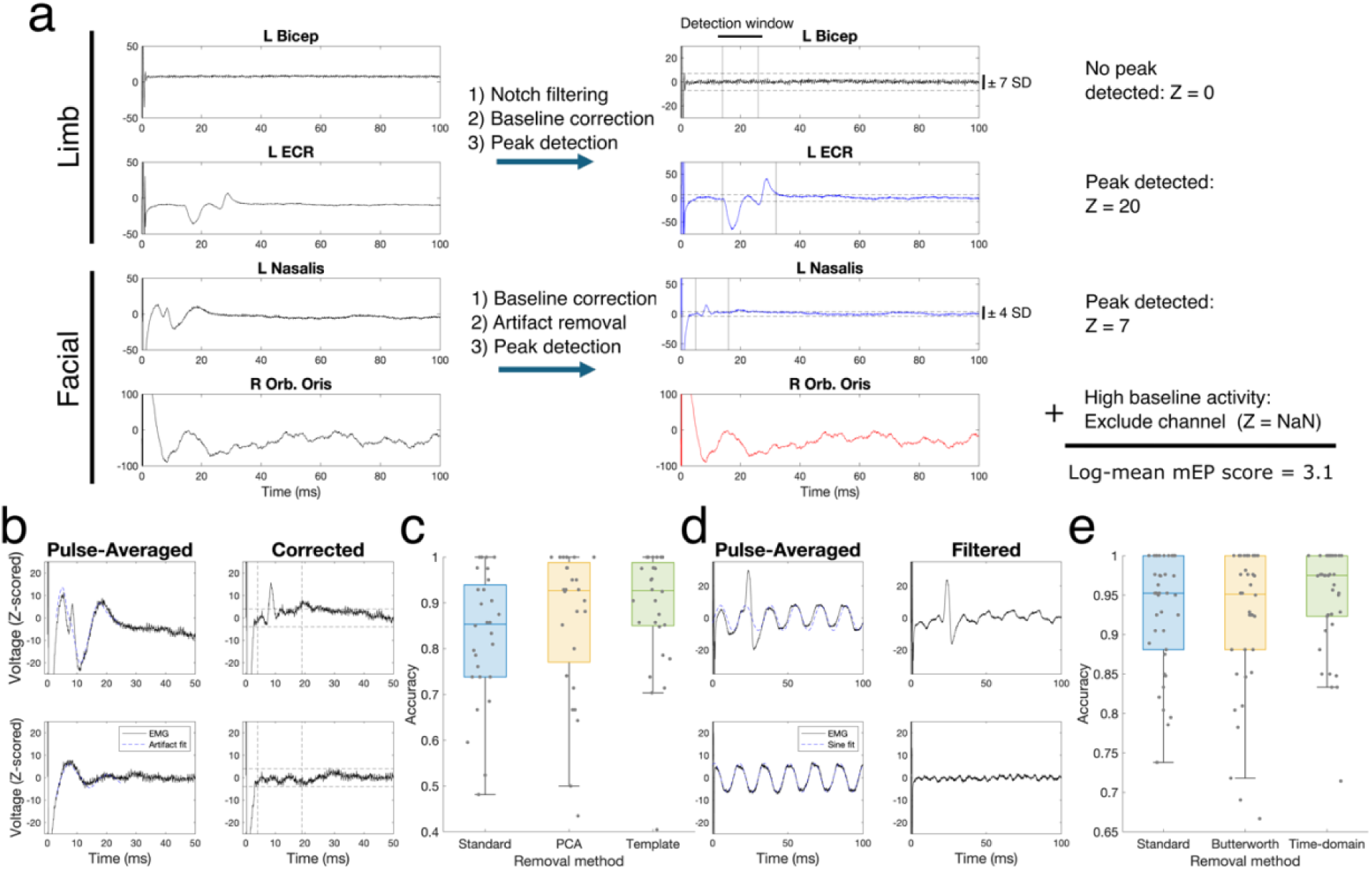
Automated motor evoked potential detection: algorithm overview and performance characterization. a) Overview of the end-to-end mEP detection algorithm for a single DBS setting. First, EMG signals are recorded from various muscles and averaged across stimulation pulses (left). Filtering, baseline correction, and latency-based peak detection steps determine whether each channel demonstrated mEP or was too noisy for accurate detection (middle). (Black: no mEP detected. Blue: mEP detected. Red: channel excluded.) Next, detected mEP peak amplitudes are normalized, averaged, and log-scaled to produce a single mEP score that quantifies the severity of capsular activation. b-e) Demonstration and quantification of critical filtering methods for mEP detection performance. b) Top: an example mEP peak (t = 9 ms) overlapping with high-magnitude stimulation artifact. Bottom: an example recording of stimulation artifact only. Right: corresponding examples after applying template-based stimulation artifact rejection. c) Detection accuracy for three artifact rejection methods (N = 28 facial muscle channels). d) Top: an example mEP peak (t = 20 ms) overlapping with 60 Hz line noise. Bottom: an example recording of line noise only Right: corresponding examples after applying a time-domain notch filter. e) Detection accuracy for three notch filter methods (N = 38 limb muscle channels).

For a subset of 9 patients (N = 66 total muscles; 28 facial muscles; 38 limb muscles; 2904 muscle/stimulation setting pairs), one investigator (SM), blinded to all algorithm development and data analysis, visually annotated the signal peaks and troughs of pulse-averaged EMG data to identify mEP responses. This dataset was used as a ground-truth evaluation set to guide the design of our prediction algorithm and to validate the accuracy of our proposed methodology. To be identified as an mEP response, the signal had to clearly deflect from the baseline and present peaks and troughs that were consistent in terms of latency and overall waveform shape within the same muscle and the same patient across multiple stimulation trials. First, mEP were identified at high stimulation amplitudes to confirm that the observed signal deflection was an mEP. High stimulation amplitudes that evoked a clear mEP then provided a visual ‘template’ to help determine whether lower amplitude stimulation settings induced mEP in the given patient and muscle.

Standard automated mEP peak detection: As a foundation for all detection strategies, we began with developing a standardized peak detection strategy which applies the following steps:

1. For a given stimulation pulse train, average the period following each stimulation pulse into a single time series event (i.e., 100 ms duration time series for a 10-Hz stimulation setting).
2. Use the ending 10-ms window of this averaged time series as an inert baseline period to compute a median and standard deviation, which is used to rectify the signal via z-score (subtract median and divide by standard deviation) followed by computing the signal absolute-value.
3. Define an mEP response by the presence of high-magnitude peaks at least N standard deviations away from the baseline median (a threshold value that we later evaluate and adjust based on the muscle category), within a given post-stimulation latency window that is customized for each muscle (e.g. 5-15 ms), and within a characteristic peak width range (peak width between 1 and 15 ms at half of maximum amplitude).

We determined the latency values used for detection in a data-driven manner. We first examined the latencies of manually annotated mEP responses to select a very wide range for detection (4-25 ms for facial muscles, 10-40 ms for limb muscles). We then ran detection across all data (N=54 leads) using these latency values and optimized detection thresholds, then examined the distributions of detected peak latencies post-hoc to derive the final prospective latency boundaries used for detection (shown in table 1). After applying peak detection, we then return the key features of the detected mEP waveform and use the greatest absolute peak height of the rectified signal to quantify mEP magnitude for a given channel.

This approach showed acceptable performance but presents limitations and is not robust to examples with high EMG noise. We amended the standard peak detection strategy by evaluating various combinations of filtering steps, particularly line noise removal, and stimulation artifact rejection (detailed in the next section) for facial muscle channels, in addition to optimizing the peak detection threshold. Filter methods are summarized below and described in detail in supplemental material.

Frequency-based filtering: we apply filtering after computing the trial-averaged signal and excluding a small period of data (2 ms) at the beginning and end of the data duration to avoid the stimulation pulses and possible edge effects of epoch-averaged data. Each filter was a second order Butterworth filter and filtering was performed using a zero-phase filtering routine to preserve signal phase.

Time-domain notch filter: we developed an alternate notch filter design that fits and subtracts a sinusoid from the data in time, rather than through convolution-based filtering. First, we compute the FFT of the 100-ms pulse-averaged time series for a given stimulation trial and compute the phase value at 60 Hz. Next, we generate a sine wave signal with unity amplitude at the computed phase, divide the data segment by this signal element-wise, and compute the median of the result. The sine wave is then multiplied by this scaling factor and subtracted from the original signal to remove the 60 Hz component (we will call this fitting technique “median-scaling” and use it again later). ^21^

### 2.4 Stimulation artifact rejection

Simulation commonly produces a post-stimulation amplifier recovery waveform artifact shaped like an exponentially damped sinusoid which can persist beyond 20 ms following the stimulation pulse (particularly for monopolar, i.e. wide field stimulation settings). This artifact waveform overlaps with the facial mEP response in both time and frequency domains, and it therefore cannot be easily separated from the mEP response waveform using conventional filtering techniques. Moreover, the shape of this artifact waveform is variable enough across individuals, channels, and stimulation trials that a one-size-fits-all approach will not be reliable – robust rejection must be performed in a data-driven manner on a trial-by-trial basis. We designed and evaluated three stimulation artifact rejection methods to improve mEP detection for facial muscles (summarized below and described comprehensively in the supplemental material).

PCA-based artifact rejection: Our first artifact rejection strategy was to compute principal component analysis (PCA) on the pulse-averaged EMG data matrix, after restricting to the time of the stimulation artifact (N channels; 2-20 ms), and then use the first principal component as a template for the stimulation artifact that can be scaled and removed from individual trials.

Template-based artifact rejection: In our second approach, we developed a library of stimulation artifact templates from held-out patient data and designed an approach to use examples from this pre-saved library to estimate, fit, and subtract the artifact from new examples (an example is shown in Fig. 2b). Because the facial mEP waveform tends to be much shorter than the full stimulation artifact duration, this method is designed to scale the template to the amplitude of only the artifact shape itself, without being influenced or distorted by a temporally overlapping mEP response waveform (especially if it is high-magnitude).

Template-PCA artifact rejection: We also implemented a hybridization that could balance the strengths of these two approaches. Before performing PCA, we identify the closest-fitting artifact template for the given example and append it as additional rows to the multi-channel data matrix (adding the appropriate number of rows is needed to double the number of channels).

### 2.5 Evaluating mEP detection

To evaluate and compare different mEP detection strategies (Fig. 2), we computed a detection accuracy values for each muscle based on the proportion of stimulation settings where the binary detection decision matched the results of manual annotation. Throughout the algorithm evaluation results (Fig. 2 and supplementary material) we report this accuracy value computed for all annotated muscles, aggregated across patients and, where applicable, separated for only facial or limb categories. On the other hand, to determine optimal detection thresholds, we use the F1 score (geometric mean of precision and recall metrics) which can be used to quantify the tradeoff between false positive and false negative detection, particularly in the case where data is very unbalanced (i.e. the majority of stimulation settings do not induce an mEP).

### 2.6 Exclusion criteria for noisy channels

For large-scale analyses, as well as real-time applications, we developed exclusion criteria based on features of the data to identify whether a given EMG channel or trial is too noisy for analysis and should be discarded (Fig. S1). Such noise could occur in real-time settings when electrodes detach from the patient’s body, poor electrode contact with the skin leads to poor signal quality, or excessive patient movement corrupts the ability to resolve a mEP response. To design these criteria, we manually identified a subset of data where at least 3 EMG channels per patient had poor-quality recording. We determined signal quality features that could distinguish good channels from bad channels. We chose the following criteria: any pulse-averaged recording that exceeds a variance during baseline period (50-100 ms) of more than 150 uV, or an average absolute value of more than 25 uV, is discarded. In later analysis, this criterion is applied separately to all channels on a trial-by-trial basis before applying mEP detection and extracting response features.

### 2.7 Computing an aggregate mEP score

To provide useful feedback for estimating side effects using mEP, it will be necessary to resolve heterogeneity across muscles and summarize this high-dimensional data into a single interpretable score that can estimate the total magnitude of capsular activation caused by a given stimulation setting – the “mEP score”. In particular, reducing the dimensionality of this information into a simplified measurement score will be critical to enable effective inference regarding how different stimulation settings selected from the high-dimensional DBS parameter space affect capsular activation, whether through manual programming or automated methods.

The mEP score computation must satisfy several properties to be a useful proxy for quantifying motor side effects. First, it should increase in value under multiple conditions: when multiple muscles have responded to stimulation vs. fewer, or whether a single muscle has responded with very high magnitude, as either property could indicate excessively increased capsular activation. Second, it should not be overly influenced by the property that some muscles may have a higher-magnitude average response than other muscles; it should provide a normalized quantification across muscles. Third, it should not be unduly influenced by a single muscle that could produce extremely high outlier response amplitudes which could indicate a saturation in side effect value and therefore eclipse the influence of other muscles on the mEP score. Fourth, it should capture the threshold-linear nature of the DBS response: many (particularly low-amplitude) stimulation parameters will not induce any capsular response and should yield a score of 0, but different stimulation settings above a certain threshold can lead to a continuously increasing activation profile. Finally, it will be helpful to have its value range be interpretable for human users. Importantly, we also design this procedure to not depend on patient-specific data distributions, so that it can be flexibly applied to real-time and/or small-data experimental settings; otherwise, this process could be simplified by e.g. normalizing muscle amplitudes based on within-patient measurements across a large number of stimulation settings.

To satisfy these conditions, we perform the following steps to compute an aggregate mEP score from multi-channel data for a given stimulation setting (overview shown in Fig. 2a):

1. First, apply exclusion criteria to identify whether any of the channels is too noisy to apply further processing.
2. Identify whether it is a facial or limb muscle; apply the optimized filtering and peak detection routine to determine whether each muscle responded.
3. For each responding channel, use the highest detected peak amplitude of the rectified signal as the mEP response amplitude. If the channel had no response, use 0.
4. For each non-zero amplitude, use pre-saved muscle amplitude distributions computed from the total patient population to normalize the amplitude of the given muscle to match the average amplitude distribution observed across all prior patients and muscles: first, subtract the detection threshold for each muscle (facial: Z = 4; limb: Z = 7); second, multiply by the pre-saved scaling factor that would adjust the interquartile range (IQR) of the given muscle to match the average IQR across muscles; third, add the limb muscle detection threshold (Z = 7).
5. Average the resulting value across (non-excluded) muscles.
6. If the result is non-zero, compute the mEP score by applying a log-transform to the result to normalize its value distribution and minimize extremely high outliers: *MEP score* = *log*2(1 + *result*)

To obtain population-level scaling factors for each muscle, we applied optimized peak detection as described above to the database of all patients. Next, we computed mEP peak amplitude distributions aggregated across all settings for each muscle, and then computed the interquartile range (difference between 25^th^ and 75^th^ percentile values) for individual muscles as well as all muscles. For each muscle, the amplitude scaling factor is the all-muscle IQR divided by the given muscle’s IQR.

Various steps of this transformation satisfy the above properties. First, using population-level interquartile ranges allow normalization across muscles in a way that respects the nonparametric and highly skewed distributions of their mEP response amplitudes across settings and patients. The final log-scaling step ensures consistent scaling even when resulting mEP values are very high and enables better interpretability of the mEP score’s operational range. Importantly, all the above steps are computationally simple and can be feasibly executed in real-time applications (approximately dozens of milliseconds in compute time to estimate the mEP score for a given stimulation setting).

The mEP score scale is designed to be interpretable: when no muscle has responded the mEP score is 0. When one muscle has produced a minimum possible mEP response, the (lowest possible) mEP score will be 1 (a single value of 8, averaged with 0-response values for all other muscles). If only 3 muscles respond, or fewer muscles have an extremely high response, the resulting value will be approximately 2 to 3. If all muscles have a high activation response, the resulting value will be approximately 6. This scale is consistent regardless of the number of muscles recorded or excluded on a trial-by-trial basis due to noise.

## 3. Results

Here, we develop and evaluate an optimized algorithmic procedure for using motor evoked potentials to quantify DBS-induced capsular activation (overview in Fig. 2a). First, we use manually annotated EMG data from 9 patients to design and evaluate a series of optimized filtering, artifact rejection, and detection steps to accurately identify mEP. Second, we analyze data from a large cohort of intraoperative experiments to study how muscles display different characteristics that can inform algorithms for mEP detection and their application, including mEP response frequency, latency, amplitude, and waveform similarity. Last, we show with individual patient data and population-level analysis that mEP respond to DBS parameter changes in a manner consistent with the anatomical relationship between lead position within the STN and the internal capsule.

### 3.1 Optimized motor evoked potential detection

We began by developing and validating an automated mEP detection approach, which quantifies whether a given muscle demonstrated an mEP for a given DBS setting. To validate accurate detection, we used manually annotated data from 9 patients. We developed a peak detection-based strategy for automatic mEP detection and varied different parameters that affect its performance, including filtering methods, detection thresholds, and stimulation artifact rejection strategies (for facial muscles only, due to their short latency, which can overlap with DBS electrical artifact). We observed that the two greatest factors affecting mEP detection are line noise and stimulation artifact.

The optimal detection algorithm required different methods for facial vs. limb muscles (Fig. 2). To improve facial muscle detection, we compared several methods to fit and remove the artifact: PCA-based rejection (removal of the 1^st^ principal component across channels); template-based rejection, which fits and removes the artifact using a pre-saved library of artifact waveforms; and template-PCA, which combines the two. We first optimized several parameters of artifact rejection (Fig. S2), finding that PCA-based rejection performs best when using only facial muscle channels for PCA. The best method to scale an artifact template for best fit to a new trial was median-scaling (see methods section 2.3), as this method was least influenced by the presence of an mEP waveform while estimating the artifact shape. We compared the best versions of each approach and found that template-based artifact rejection performed best, particularly in the worst-case (from a lower bound of 48% to 70% accuracy) (Fig. 2b-c). Notch filtering did not improve facial muscle detection alone (Fig. S3), nor in conjunction with artifact rejection (Fig. S4).

Limb muscle detection was best improved by a time-domain notch filtering strategy (Fig. 2d-e), which fits and subtracts a 60-Hz sinusoid from the signal (instead of a conventional Butterworth filter), improving both median (95% to 98%) and worst-case detection accuracy (from 75% to 84%). Lastly, we swept the peak detection threshold and chose 4 standard deviations from baseline for facial muscles, and 7 standard deviations for limb muscles (Fig. S5).

### 3.2 mEP response characteristics vs. muscle type

Next, we applied optimized mEP detection to a large-scale database of intraoperative recordings (N=54 DBS leads) to study how characteristics of the mEP response may differ between muscles, and how such characteristics may guide the use of mEP for measuring capsular activation (Fig. 3). We first examined the response frequency of different muscles, asking whether some muscles are preferentially activated by stimulation more frequently than others (Fig. 3a) – a property that could guide where EMG electrodes should be best placed to estimate activation for future patients. We quantified the proportion of stimulation settings where each muscle displayed above-threshold activation, out of all stimulation settings that produced activation in at least one muscle. We found that facial muscles responded more frequently than limb muscles (p = 5.68e-26; generalized linear mixed effects model using binomial distribution, with random effects for lead and muscle; N = 461 facial muscle responses and 72 limb muscle responses), with more modest differences within muscle groups. Despite this population-level trend, 16 out of 54 DBS leads featured a higher proportion of limb responses than facial responses.

**Figure 3.**
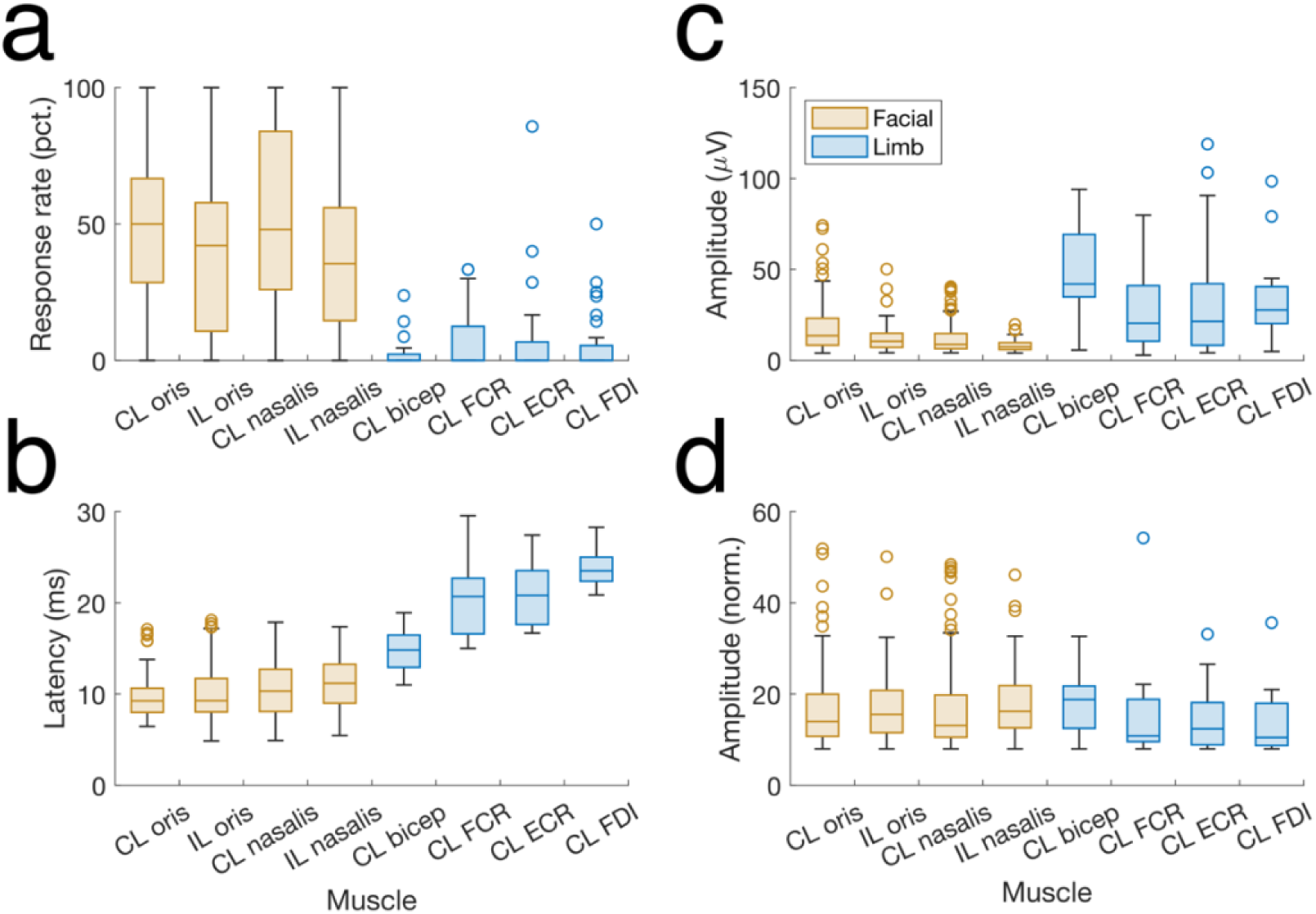
Motor evoked potential response characteristics by muscle type. a) Percentage of stimulation settings where a given muscle responded with mEP, out of stimulation settings where at least one muscle responded. (facial vs limb: p = 5.68e-26; generalized linear mixed effects model using binomial distribution, with random effect for lead) b) mEP peak latency for different muscles (sorted from lowest to highest latency response). (facial vs limb: p = 7.37e-75; linear mixed effects model vs. lead) c) Detected mEP response amplitudes across patients for various muscles. (facial vs limb: p = 0.022; linear mixed effects model with random effect vs. lead) d) Normalized amplitude distributions after applying signal baseline correction, z-scoring, and IQR normalization on each muscle group to match the average across-muscle IQR. CL: contralateral to the stimulated hemisphere. IL: ipsilateral to the stimulated hemisphere. N = 461 facial muscle responses; 72 limb muscle responses.

Second, we found that mEP response latencies (temporal delay between the stimulation pulse and detected mEP activation peak) are highly stereotyped across muscles and consistent with the path length between the site of capsular activation and muscle location – an expected physiologic property that guided the development of our automated detection strategy. We observed comparable response latencies across facial muscles, with the clearest delineation across limb muscles: the anatomically closest muscle group (bicep) featured the shortest peak latency, and the anatomically furthest group (FDI, which controls the index finger) featured longest latency. (facial vs limb: p = 7.37e-75; linear mixed effects model, with random effects for lead and muscle)

Next, we similarly studied the characteristics of detected mEP peak amplitudes (aggregated across all patients and stimulation settings) and their variability across muscle type (Fig. 4c). mEP amplitude distributions varied substantially across muscles in the median amplitude, variability, and degree of skewness. Facial muscles including nasalis featured smaller responses. Most muscles featured high upper bounds indicating a skewed distribution with a substantial number of high-magnitude outliers. To develop a single mEP biomarker score, we designed an IQR (inter-quartile range) normalization scheme that was successful in minimizing variability between the non-normal mEP amplitude distributions across muscles (shown in Fig. 4d).

**Figure 4.**
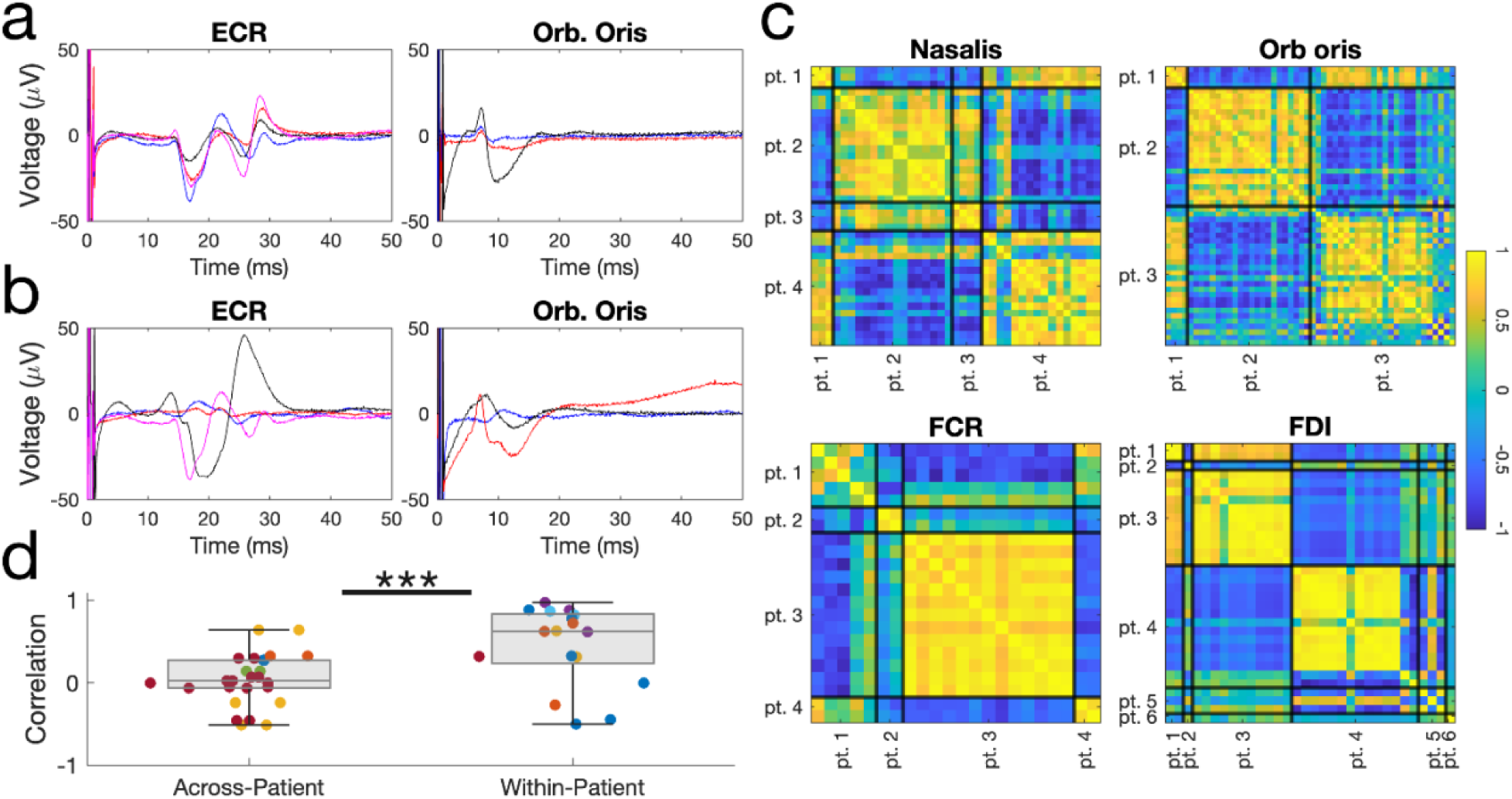
Motor evoked potential waveform shapes are highly correlated within-patient and within-muscle. a) Representative recordings from different stimulation settings within the same muscle and patient, which share similar features and response shape. b) Example recordings from the same muscle but different patients, which vary in response shape. c) Correlation matrices quantifying mEP waveform similarity (Pearson’s correlation coefficient between waveforms of different stimulation trials, restricted to the expected muscle peak latency) across stimulation settings from several example patients, for the same muscle type. Black lines: boundaries separating sets of stimulation settings that were collected from different patients. d) Waveform correlation coefficient averaged across pairs of mEP waveforms for different stimulation settings, separated by within-patient vs. across-patient comparisons. Colors correspond to different muscles. (across-patient vs. within-patient: p = 0.0032; Wilcoxon rank-sum test; N = 26 vs. 17 comparison pairs)

### 3.4 mEP response waveforms are correlated within-subject and within-muscle

Next, we found that the mEP waveform shape is highly stereotyped across stimulation settings for the same patient and same muscle (Fig. 4). However, its shape is highly variable when recording from the same muscle in different patients. To study this property, we used the evaluation set of held-out patient data, identified all stimulation settings where a given muscle responded, appended the pulse-averaged EMG recordings into a matrix (segmented for only the selected latency window used for detection on the given muscle), and computed a correlation matrix quantifying waveform similarity for all pairs of stimulation settings (Fig. 4c). We observed that this correlation matrix had visually apparent block-diagonal structure that aligned to trial boundaries between patients and was consistent for different muscles. Waveform pairs from trials within the same patient and muscle had a higher correlation coefficient (median of 0.62) than waveform pairs selected from different patients (median 0.02) (p = 0.0032; Wilcoxon rank-sum test; N = 26 vs. 17 pairs). Though our detection method did not use this property, high within-patient correlation could be exploited as a future direction for improving automated mEP detection. Additionally, this result validates a key assumption which was used to provide the manual ground-truth data set used to assess automated mEP detection. Last, this property provides a useful heuristic for researchers and clinicians to clearly differentiate physiological mEP responses from various forms of system noise that can arise in surface EMG recordings.

### 3.5 Motor evoked potentials respond to changes in DBS amplitude and contact configuration

Last, we examined how DBS parameters, including amplitude and contact, affect mEP and whether the profile of mEP modulation by DBS parameters is consistent with the expected relationship between DBS contacts, the STN, and capsular anatomy. Fig. 5 visualizes how various DBS parameters, including contact configuration and amplitude, increase the mEP score (Fig. 5a) alongside pulse-averaged recordings from several muscles (Fig. 5b) in an example patient. This example shows a plausible capsular response profile to DBS, in which all contacts exhibit threshold-like recruitment curves with the thresholds themselves being contact-dependent. This example also demonstrates the utility of the mEP score, which summates high-dimensional recordings into a single value that better enables comparison and inference across different settings selected from the high-dimensional DBS parameter space.

**Figure 5.**
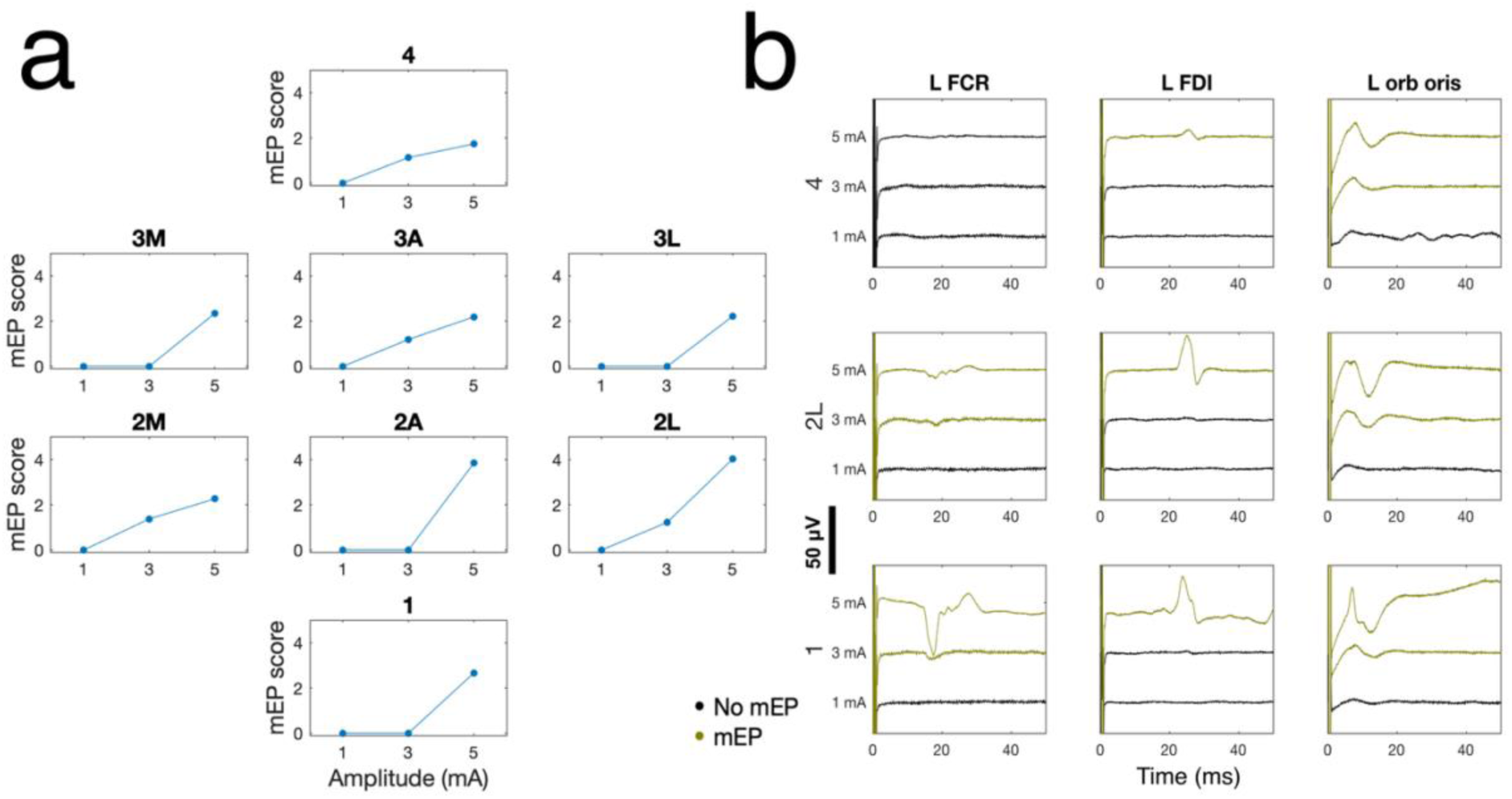
DBS parameters modulate motor evoked potentials in an example patient. a) mEP score vs. stimulation amplitude with DBS targeting right STN, plotted separately for different monopolar contact configurations (all settings at 60 us pulse width). Subplots are arranged by geometric position of the stimulated contact along the DBS lead. M: posteromedial; A: anterior; L: posterolateral; 1: most ventral level; 4: most dorsal level. b) Pulse-averaged EMG recordings corresponding to different DBS amplitudes, visualized for three monopolar contact configurations and three recorded muscles.

Next, we quantified how DBS parameters modulate mEP at the population level. We first found that DBS amplitude increases both the peak amplitude of detected mEP (Fig. 5a; p = 4.68e-03; linear mixed effects model; N = 530 responses; random effect vs. patient and muscle type) and proportion of responding muscles (Fig. 5b; p = 6.20e-10; N = 1379 settings; linear mixed effects model; random effect vs. patient), providing justification for the inclusion of both factors in designing a single biomarker score. Next, we examined how DBS contact configuration and amplitude modulate mEP. First, we grouped DBS amplitudes into low (0-2 mA), medium (2-4 mA), and high (4-6 mA) groups and plotted a recruitment curve of mEP score vs. amplitude for different contacts. In visualizing the DBS response this way, a few properties are expected. First, we observed no activation for the vast majority of low-amplitude settings. Second, we observed a higher mEP score for monopolar stimulation using contacts that are typically located closest to the internal capsule, including level 1, level 4, and lateral segments 2L and 3L,^22^ compared to other contacts (Fig. 6e) (p = 0.032; N = 133 near IC settings vs. 167 other settings; linear mixed effects model with random effect vs. lead). Despite high variability across patients – as expected, given variability in local anatomy and surgical targeting – these recruitment curve characteristics replicate expected properties of STN-capsular anatomy and suggest that mEP can provide a useful biomarker for quantifying DBS-induced capsular activation.

**Figure 6.**
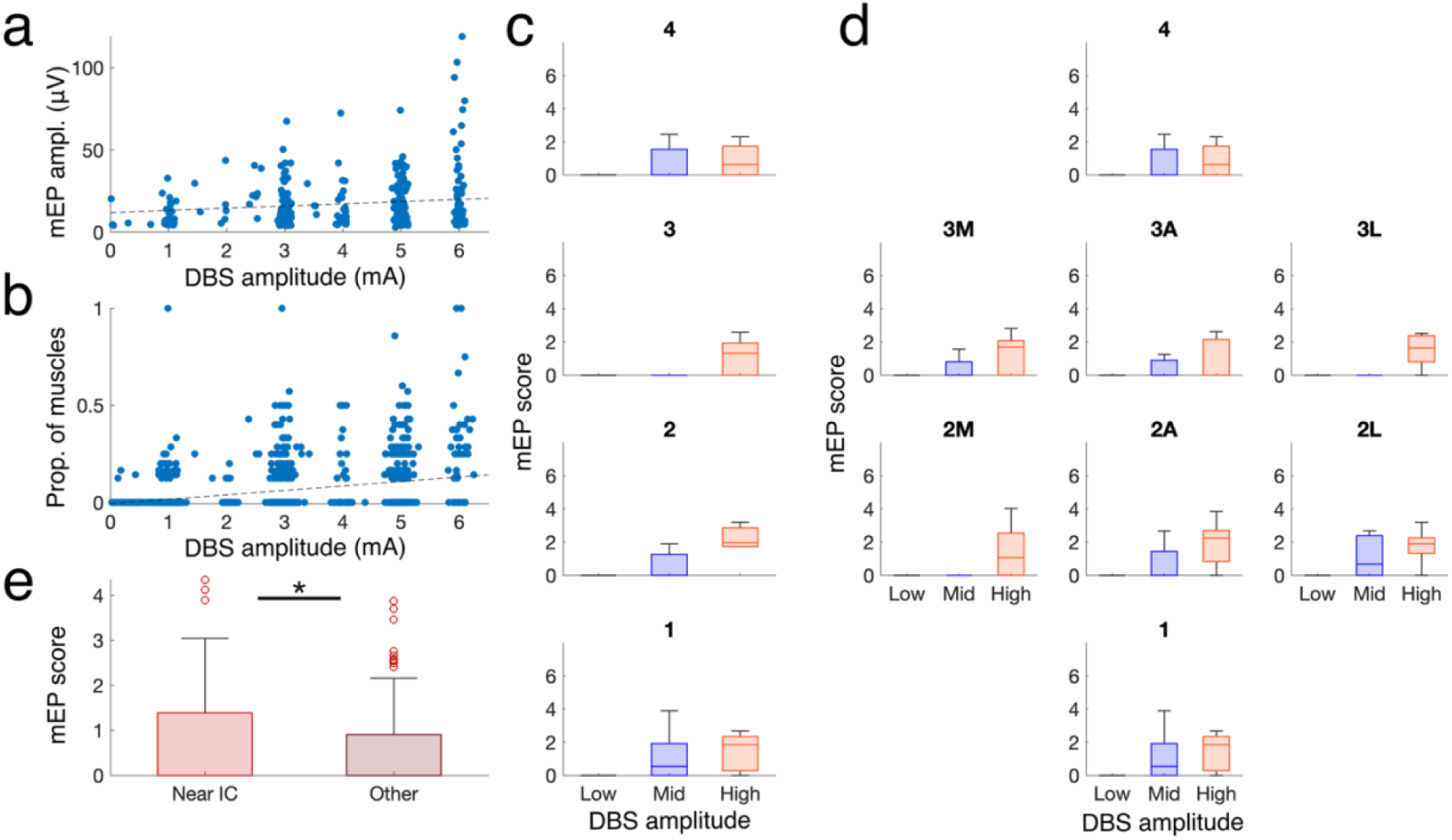
Motor evoked potentials respond to changes in DBS amplitude and contact configuration. a) mEP peak amplitude vs. stimulation amplitude. (p = 4.68e-03; linear mixed effects model; N = 530 responses; random effect vs. N=54 leads) b) Proportion of muscles with a detected mEP response vs. stimulation amplitude (p = 6.20e-10; linear mixed effects model; N = 1379 settings; random effect vs. lead). c) mEP score vs. stimulation amplitude and full-ring monopolar contact configurations. d) mEP score vs. stimulation amplitude and segmented monopolar contact configurations (shown for N=11 patients where stimulation was performed at all segments and amplitudes). Low: 0-2 mA; Mid: 2-4 mA; High: > 4 mA. Subplot orientation corresponds to the geometry of the DBS device: e.g. ‘1’ indicates stimulation delivered at the most ventral contact; ‘4’ at the most dorsal contact; etc. Segment orientation: A = anterior segment; M = medial segment; L = lateral segment. e) mEP score of monopolar contacts at 3 mA amplitude, grouped by expected anatomical proximity to the internal capsule. Near IC: monopolar stimulation at contacts 1, 2L, 3L, and 4. Other: monopolar stimulation at all other contacts. (p = 0.032; linear mixed effects model; N = 133 near IC settings vs. 167 other settings; random effect vs. lead)

## 4. Discussion

### Conclusion

In this study, we demonstrated an automated procedure for estimating capsular activation using mEP as an objective biomarker for DBS-induced motor side effects. We first developed and validated a signal processing algorithm for automated detection, then designed a single mEP activation score that summates multi-muscle responses to quantify the effect of a given DBS setting. Next, we quantified the response frequency of different muscles, showed that different muscles have stereotyped and anatomically consistent response latencies, and showed that waveform shapes are consistent within-muscle and within-patient. Last, we showed that mEP responds to stimulation parameters in a manner that is consistent with the spatial profile of capsular activation by STN DBS. This approach could provide objective feedback to improve the precision and speed of DBS programming and surgical targeting, and if paired with other biomarkers for DBS-induced symptom relief, could pave the way for full automation of programming.

### Implications

The results of this study could inform a complete experimental protocol for using mEP as a biomarker to estimate motor side effects. First, our results inform where multiple EMG channels should be placed to most likely capture activation based on their response probability: prioritizing facial muscles where feasible, then forearm muscles (ECR, FCR), then FDI, then biceps (however, this relationship was not consistent across all patients, necessitating multi-muscle recording for detection of personalized activation profiles). Second, we provide an end-to-end algorithmic procedure that can estimate capsule activation for a given stimulation setting to provide interpretable biomarker feedback, enabling more scalable comparison of DBS settings.

### Prior work

Although a range of algorithmic approaches have been developed for detecting mEP (mainly in the context of transcranial magnetic stimulation), our work is novel in the context of DBS, which poses some distinct challenges. Prior TMS-focused studies have employed techniques such as the squared hard threshold estimator,^23^ derivative-based methods for estimating mEP latency,^24^ and traditional filtering and peak detection algorithms for identifying response amplitudes.^25^ While our approach incorporates some strategies for peak detection and filtering that are similar to prior work, several characteristics unique to DBS-evoked responses necessitated different methods – namely, the specific waveform characteristics of stimulation artifacts and their high magnitude relative to the lower DBS mEP amplitude. Additionally, our approach reflects differences in goals between TMS and DBS: while many TMS-mEP studies have focused only on extracting amplitude and latency features, we place a greater emphasis on accurately identifying whether mEP were present, as navigating the boundary between side-effects and the lack thereof is crucial for DBS.

### Future directions

While our method was able to achieve a median detection accuracy above 90%, its performance suffered in some challenging scenarios – for example, low-amplitude facial muscles coupled with high noise and large stimulation artifact. Here, we investigated the extent to which standard signal processing methods could accurately resolve mEP. However, future work could consider more advanced detection strategies beyond peak detection that leverage unique properties of the mEP data to approach 100% accuracy. Based on our result demonstrating within-muscle waveform similarity, one could derive a response template from high-activation DBS settings to guide a template-based filtering or detection approach, or apply correlation-based detection approaches to detect responses based on similarity across stimulation trials.^26–28^ However, we did not investigate such approaches due to the added complexity such methods introduce in data-constrained and real-time settings, where a clear response template or multiple responding trials are not guaranteed to appear. Another direction could use waveform-based pattern recognition algorithms such as convolutional neural networks, which may better discriminate between mEPs and stimulation artifact than peak detection.^29^ Detection strategies could also be augmented to incorporate user-informed heuristics; for example, using the DBS parameters of the given stimulation trial or examining whether multiple muscles responded to help assess whether a mEP response was physiologically plausible.

### mEP and side effects

Importantly, in this study we did not directly evaluate mEP as a biomarker of side effects using patient-reported side effect data. This relationship has already demonstrated in prior work. Our previous study identified a correlation between DBS amplitude thresholds that induce patient-reported side effects and mEP in the context of post-surgical programming.^17^ Further work has shown that mEP increase with reduced electrode distance to the internal capsule and vary with side-effect thresholds.^18,19^ As an alternative to our intraoperative experiment design, EMG recordings performed in the clinic could enable more comprehensive testing of stimulation settings and allow more time to assess EMG recording quality, which may be better suited to directly address the clinical utility of mEP. Future work could address unanswered questions, including prospectively evaluating side-effect severity of parameters selected at the mEP threshold, studying whether muscle-specific activation predicts the spatial location of side effects, and to what degree mEP response magnitude corresponds to side effect perceptibility and severity.

### Significance

Our proposal of mEP as a biomarker of side effects complements a growing literature of techniques that could be used to automated DBS programming,^16^ including biomarkers for symptom relief (such as subthalamic beta power,^30^ gamma power,^31,32^ and DBS evoked potentials^33,34^) and algorithms that can automatically search for optimal settings based on biomarker values (such as Bayesian optimization,^35^ its extensions,^36–39^ and related methods^40–42^). By incorporating automated mEP detection into platforms for real-time stimulation and recording, clinicians could obtain faster and more objective feedback regarding side effects to improve the efficiency and precision of DBS programming and surgical targeting. The ability to perform mEP recordings under anesthesia could enable motor side-effect feedback during asleep surgery, which could aid the precision of surgical targeting.^18^ Additionally, while we focused on the STN in this study, our methodology for mEP detection should be readily applicable to other DBS targets that can cause motor side-effects through capsular activation, including the globus pallidus internus and ventral intermediate nucleus of the thalamus.^43^ Finally, mEP could be used to further refine and guide the development of other technologies for DBS, including computational modeling and image-guided parameter selection tools.^44,45^ Long-term, such technical advancements could improve the reliability of DBS treatment and expand its availability to aid more patients with Parkinson’s disease and other movement disorders.

### Limitations

A primary limitation in this study comes from the constraints of retrospective analysis using data collected from time-limited intraoperative research. As a result, only a small set of stimulation settings could be tested for each patient, which also differed in which amplitudes and/or contacts were tested across patients. Second, the intraoperative research environment can result in noisier EMG recording. Although we incorporated automatic trial-level exclusion criteria to account for these challenges, this property could result in some trials or patients having higher variability in the mEP score if multiple trials or channels were excluded. Additionally, the use of low-frequency stimulation to predict clinical DBS effects assumes that the spatial elements of activated tissue are consistent between single pulse and high-frequency stimulation. This expectation is supported by computational modeling studies of DBS^46,47^ and experimental studies that demonstrated a relationship between biomarkers measured at low frequency and side-effects at high frequency,^17^ but may need to be further tested experimentally. Last, we did not quantify how different lead placement across patients affects the mEP response, a property that introduces high variability in population-level analysis regarding the effects of DBS parameters. We consider this question outside the scope of our study, as this relationship has been investigated previously and there are many implementational complexities underlying the analysis of imaging and estimation of pathway activation by DBS that merit a standalone investigation.

## 5. Data Availability Statement

Code that support the findings of this study are openly available at the following URL/DOI: https://github.com/ericrcole/mep_detection/tree/main. The data that support the findings of this study are available upon reasonable request from the corresponding author.

## 6. Acknowledgements

This work was supported by NINDS (5R01NS125143) and the McCamish Foundation Parkinson’s Disease Innovation Program.

## 7. Author Contributions

E.R.C. led data analysis and manuscript preparation. E.O. aided in conceptualizing the study, developed initial methodologies and performed experiments. S.B.B., F.I., J.T.W., N.A.Y., and R.E.G. performed experiments and data collection. Y.H. contributed to data analysis. S.M. conceptualized and supervised the study, performed experiments and prepared the manuscript.

## Notes

### Competing Interest Statement

The authors have declared no competing interest.

### Funding Statement

This work was supported by NINDS (5R01NS125143) and the McCamish Foundation Parkinsons Disease Innovation Program.

### Author Declarations

The IRB of Emory University gave ethical approval for this work.

