## Supplemental Material for "An algorithmic procedure for measuring deep brain stimulation-induced capsular activation using motor evoked potentials"

### SUPPLEMENTARY MATERIAL

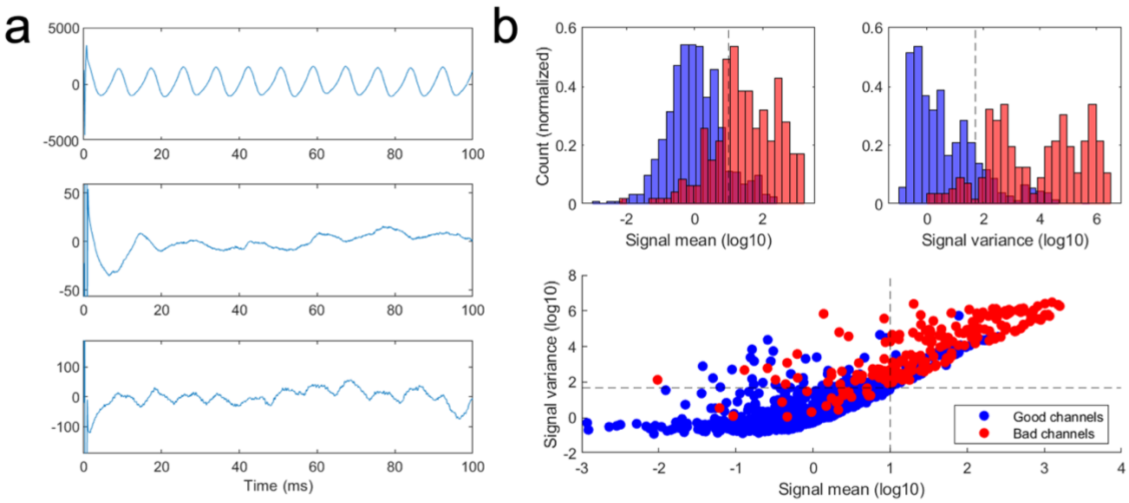


**Fig. S1: Exclusion criteria for motor evoked potential detection.** a) Examples of unusable pulse-averaged EMG recordings, which can include very high-magnitude 60 Hz line noise (and/or its harmonics) or highly fluctuating baseline activity indicating possible electrode detachment. b) Statistics computed during baseline (50-100 ms post-pulse) can distinguish good vs. bad channels. We manually identified channels in 3 patients that were visually deemed unsuitable for manual mEP annotation, and aggregated data for all DBS settings from these patients into 'Good channel' and 'Bad channel' groups. Black dashed lines: manually chosen feature thresholds used to automatically exclude channels for population analysis and other automatic detection use cases (variance > 50 uV or absolute mean >10 uV). Because we could detect mEP responses across any of 8 muscles for each patient, we chose low feature thresholds which could make channel exclusion more likely and prioritize reducing the possible influence of false positive detections that may result from high noise. However, thresholds could be made more conservative for different experimental settings based on other relevant criteria.

*Filtering methods*

Frequency-based filtering: a first-pass approach would be to use standardized filtering (e.g. a convolution-based Butterworth filter) prior to pulse-averaging and applying peak detection. However, convolution with the sharp stimulation artifact pulse during filtering can introduce large artifacts into the data to make detection worse. We instead apply filtering after computing the trial-averaged signal and excluding a small period of data (2 ms) at the beginning and end of the data duration to avoid the stimulation pulses and possible edge effects of epoch-averaged data. Moreover, filtering (particularly a 60-Hz notch filter) could remove frequency components of the mEP waveform itself and worsen detection accuracy by distorting its shape. Each filter was second order and filtering was performed using a forward and reverse filtering routine to preserve signal phase.

Time-domain notch filter: We developed an alternate notch filter design that fits and subtracts a sinusoid from the data in time, rather than through convolution-based filtering. First, we compute the FFT of the 100-ms pulse-averaged time series for a given stimulation trial and compute the phase value at 60 Hz. Next, we generate a sine wave signal with unity amplitude at the computed phase, divide the data segment by this signal element-wise, and compute the median of the result. The sine wave is then multiplied by this scaling factor and subtracted from the original signal to remove the 60 Hz component (we will call this fitting technique "median-scaling" and use it again later). This technique has several benefits that are suitable for the structure of this problem. It leverages the property that the mEP response tends to be temporally sparse and short-duration; by scaling based on median fit, the magnitude of the 60-Hz component is primarily fit to data outside the mEP response where there is only line noise, and the shape of an mEP response that may be present is preserved. This method also assumes that the phase and magnitude of the sinusoidal component are consistent over the trial duration, which is more appropriate for the 100-ms epoch-averaged trial (only 6 cycles within 100 ms).

*Stimulation artifact removal methods*

PCA-based artifact rejection: We first observed that cross-channel data can be used to differentiate the stimulation artifact from mEP response: the shape of the artifact will often take a similar form across different facial muscle channels, whereas the mEP response on different channels will occur at variable latencies and with differing waveform shapes. Our first artifact rejection strategy was to compute principal component analysis (PCA) on the pulse-averaged EMG data matrix, after restricting to the time of the stimulation artifact (N channels; 2-20 ms) and use the first principal component as a template for the stimulation artifact that can be scaled and removed from individual trials. We evaluated multiple design criteria that could affect the utility of this approach, including which data is used as the basis for PCA to estimate the artifact (all EMG channels, only facial channels, or appending channels from simultaneous DBS LFP recording) and what method is used to scale the template to individual trials and channels (removal of the 1$^{st}$ PC, fit by minimizing Euclidean distance, or median-scaling).

Template-based artifact rejection: In our second approach, we developed a library of stimulation artifact templates from held-out patient data and designed an approach to use examples from this pre-saved library to estimate, fit, and subtract the artifact from new examples (an example is shown in Fig. 2b). To develop this template library, another blinded researcher (YH) visualized pulse-averaged EMG data from facial muscles in the remainder of the complete dataset (i.e., patients separate from the 9-patient evaluation dataset) to identify examples that have clearly identifiable stimulation artifact with no physiological mEP response. 192 example trials were identified and saved. Next, we applied baseline correction (as described in 2.3) to each example, applied a Savitsky-Golay filter (2$^{nd}$ order polynomial smoothing using a 51-sample window size) to reduce high-frequency noise, and concatenated the data matrix with a negated copy of the matrix to produce the final template library (384 examples). To remove the artifact prospectively from a given recording: first, we compute the Pearson's correlation coefficient between the new recording waveform and all examples from the template library to find the template that most closely matches the new example in waveform shape (restricted to a pre-selected time over which to remove the stimulation artifact, 2-20 ms). Then, we use the median-scaling method described previously to multiply and rescale the template for closest fit to the new example. Because the facial mEP waveform tends to be much shorter than the full stimulation artifact duration, this method is designed to scale the template to the amplitude of only the artifact shape itself, without being influenced or distorted by a temporally overlapping mEP response waveform (especially if it is high-magnitude).

Template-PCA artifact rejection: The two aforementioned methods feature complementary designs. The PCA method can reliably estimate the artifact shape in a data-driven manner, but it could also remove part of the mEP response if an existing mEP waveform is sufficiently high-magnitude to appear in the first principal component of the data, or multiple channels happen to have an mEP response that is similar in latency and waveform shape. Conversely, template-based rejection is designed to ensure that the artifact template does not overfit to incorporate any mEP response, but may underfit to the specific artifact shape of new individual trials, particularly if they are out-of-distribution with respect to the template library. We therefore also implemented a hybridization that could balance the strengths of these two approaches. This method is an extension of PCA-based rejection where, before performing PCA, we identify the closest-fitting artifact template for the given example and append it as additional rows to the multi-channel data matrix (adding whatever number of rows is needed to double the number of channels). This approach might therefore better estimate the artifact on a trial-by-trial basis by reducing the potential to overfit to mEP response waveforms while maintaining trial-specific, data-driven artifact estimation.

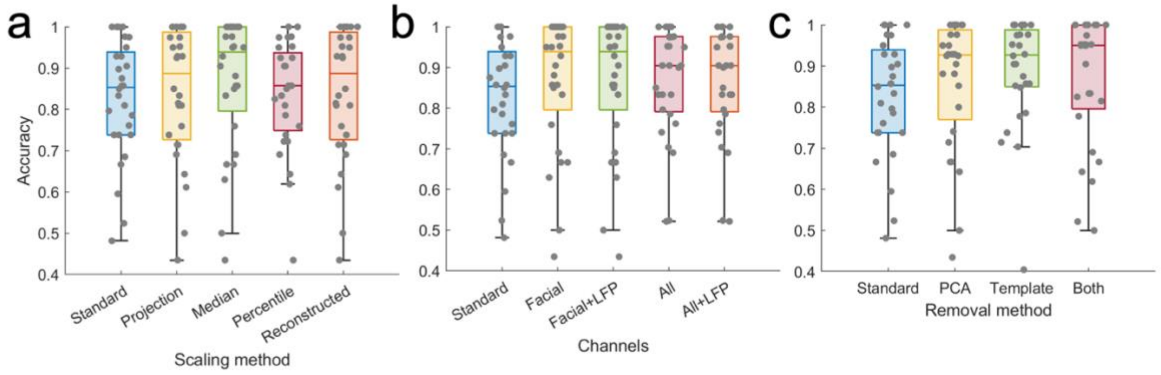

**Fig. S2: Detection accuracy for various stimulation artifact rejection parameters.** a) Comparison of different data sources used as the basis for fitting the stimulation artifact with PCA. Standard: basic latency-based peak detection without artifact rejection. Facial: facial muscle channels only. All: both facial and limb muscle channels. +LFP: intraoperative DBS-LFP recordings from the STN are concatenated with EMG data before applying PCA. b) Comparison of key artifact rejection methods (both: template-PCA method). c) Accuracy comparison of different scaling methods used to fit an acquired template waveform to each given trial before subtracting. Projection: least-squares fit minimizing Euclidean distance between the template and data. Median: median-scaling method. Percentile: an equivalent to the median scaling method, instead using the 20$^{th}$ percentile of the elementwise dividend. Reconstructed: each trial is reconstructed after discarding the 1$^{st}$ principal component.

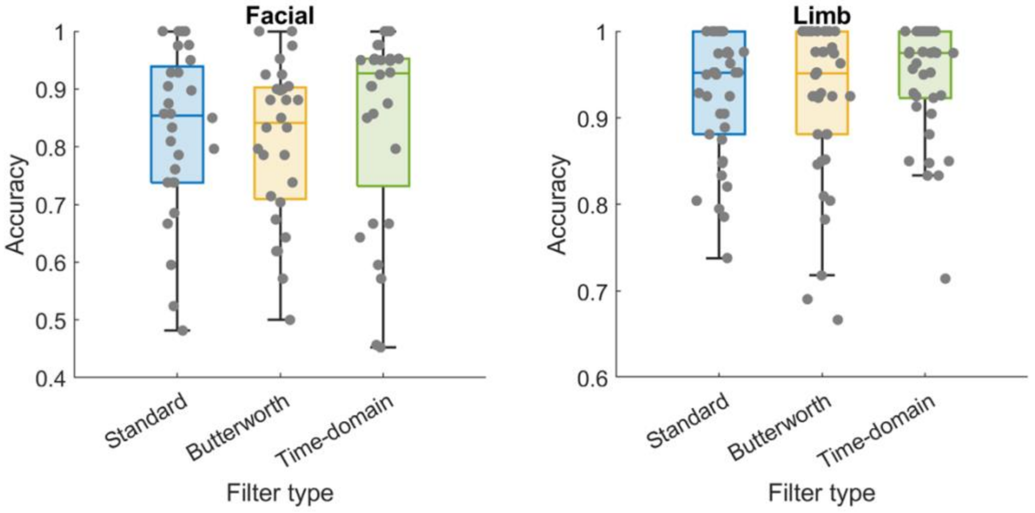


**Fig. S3: Detection accuracy for various notch filter designs.** Left = facial muscles; right = limb muscles. Butterworth: Butterworth notch filter, as described in methods. Time-domain: time-domain notch filter.

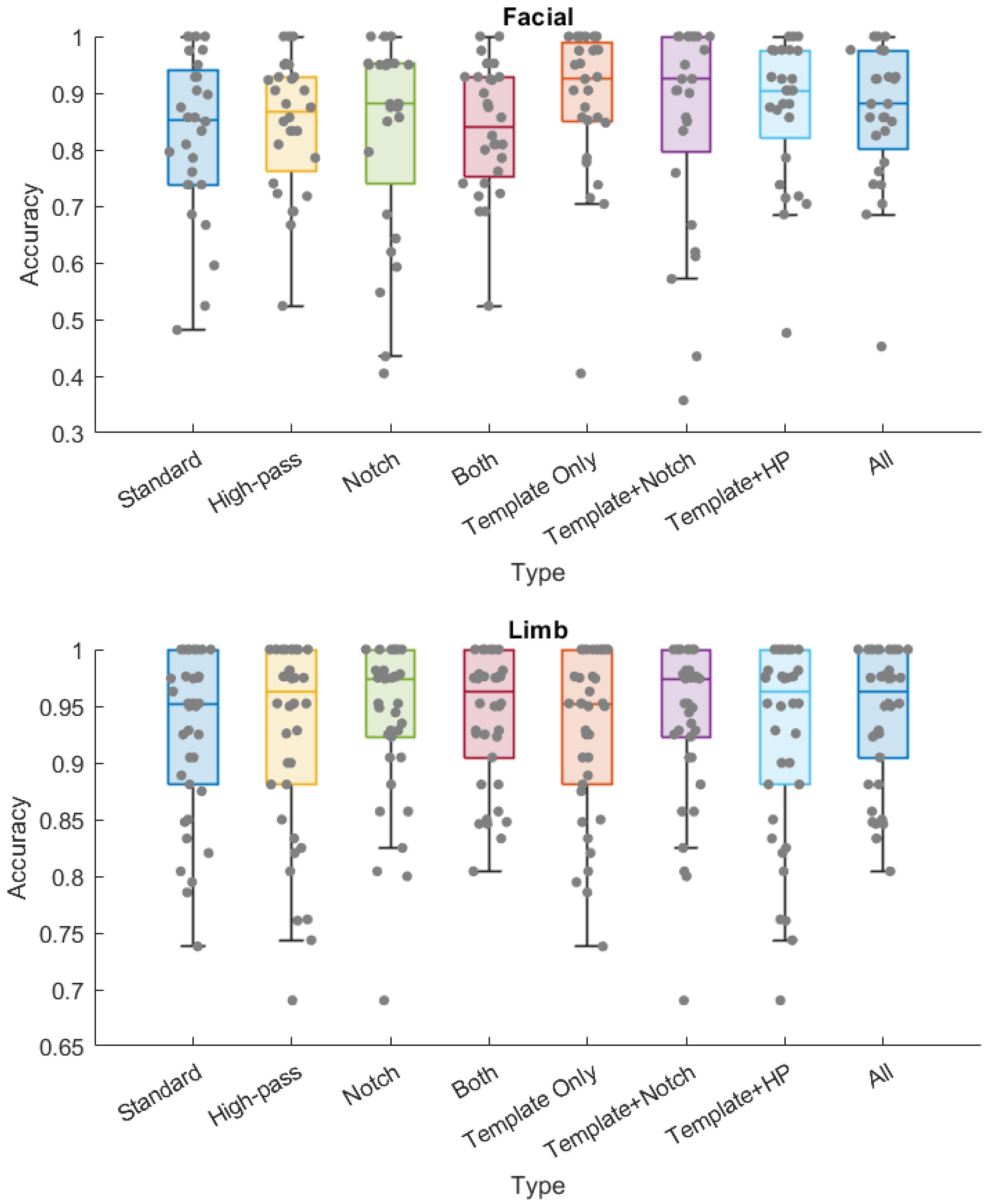


**Fig. S4: Detection accuracy for combinations of artifact rejection and filtering methods.** High-pass (HP): 30 Hz high-pass filter. Notch: time-domain notch filter routine. Template: template-based artifact rejection.

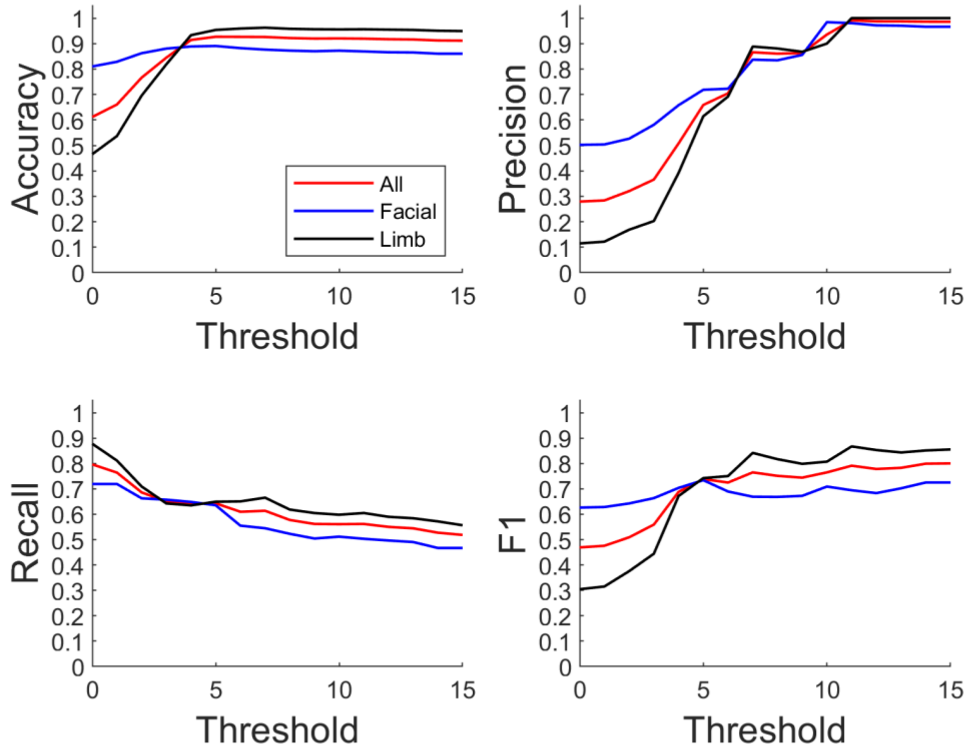


**Fig. S5: MEP detection precision-recall analysis vs. detection threshold.** Each metric is computed by comparing the number of equally predicted stimulation settings between automated detection at the given threshold vs. manual annotation. Each curve shows the mean value across the given muscle group. Threshold: minimum detected peak height, in number of standard deviations above baseline activity.

Accuracy: $(TP + TN)/(TP + TN + FP + FN)$

Precision: $TP/(TP + FP)$

Recall: $TP/(TP + FN)$

F1: $2 * (Precision * Recall)/(Precision + Recall)$

TP: true positive count; TN: true negative count; FP: false positive count; FN: false negative count.
